# A new surveillance landscape for seasonal influenza?: Comparing lab-confirmed influenza hospitalizations with other syndromic and surveillance data sources for the state of California

**DOI:** 10.1101/2025.10.28.25338991

**Authors:** Lauren A. White, Monica Sun, Elisabeth Burnor, Erin L. Murray, Cora Hoover, Christina Penton, Nancy J. Li, Floria Chi, Cassandra O. Schember, Tomás M. León

## Abstract

**Background:** During the COVID-19 pandemic, U.S. federal hospitalization surveillance systems provided a new data source for lab-confirmed influenza hospital admissions; these data were important for monitoring hospital capacity for combined respiratory virus pathogen burden and served as a new forecasting target for seasonal influenza forecasting efforts.

**Methods:** For influenza surveillance in the state of California, from February 2, 2022 through May 31, 2025, we explored: (1) how lab-confirmed hospital admissions correlated with other syndromic and sentinel surveillance data; (2) which signals could serve as a potential leading input for predicting hospital admissions; and (3) how these relationships varied across respiratory virus seasons and geography.

**Results:** Despite varying across seasons and regions, influenza surveillance data sources in California were strongly correlated with laboratory-confirmed influenza hospitalizations (Spearman’s ρ ≥ 0.8). These correlations generally strengthened through time from the 2021-2022 season to the most recent 2024-2025 season, especially for death, influenza-like illness, wastewater, and clinical lab data. Most of these data sources neither consistently led nor lagged hospital admissions across all four seasons, but electronic laboratory reporting provided consistent leads of one to two weeks relative to the admissions signal across all four seasons (Spearman’s ρ ≥ 0.89). Principal component analysis suggests that 93% of the variation in data signals can be explained by a single axis.

**Conclusions:** Influenza surveillance data sources have inherent trade-offs in geographic coverage, temporal resolution, and reporting frequency. Understanding the relationship between different data sources will inform future predictions of influenza burden, including forecasting and scenario modeling.

**Key Messages:**

- Influenza surveillance data sources in California were strongly correlated with new laboratory-confirmed influenza hospitalizations data from the National Hospital Safety Network (NHSN), and these correlations strengthened through time from the 2021-2022 to the most recent 2024- 2025 season.
- Although most data sources did not provide a clear leading or lagging signal, electronic laboratory reporting results provided consistent leads of one to two weeks relative to laboratory-confirmed influenza hospital admissions.
- This analysis demonstrates that existing influenza surveillance and sentinel data sources correspond well with the new NHSN data, which will be important for ongoing surveillance and public health action during future respiratory virus seasons.

## Introduction

Prior to the COVID-19 pandemic, seasonal influenza was the major driver of respiratory virus disease burden in the United States, with significant effort dedicated to tracking, measuring, forecasting, and estimating influenza activity. However, the United States lacked a unified system for tracking all influenza inpatient hospitalizations, a key metric for healthcare system capacity. Laboratory-confirmed hospitalizations from the Influenza Hospitalization Surveillance Network (FluSurv-NET) were used for influenza burden estimation but had limited geographic coverage,^1^ representing only a few states across the United States and only certain counties within those states.^2^ The Centers for Disease Control and Prevention (CDC)’s collaborative FluSight Forecasting Challenge began in 2013 and focused on outpatient influenza-like illness (ILI) as the target metric.^3,4^

The COVID-19 pandemic prompted a new influenza surveillance and forecasting paradigm with the advent of mandatory reporting of laboratory-confirmed influenza hospital admissions through the U.S. Health and Human Services (HHS) Protect and later National Healthcare Safety Network (NHSN) platforms. This development was important, as pandemic-driven changes in healthcare seeking behavior and testing patterns complicated the reliability and interpretability of the ILI metric, since COVID-19 was often conflated with influenza at the beginning of the pandemic. Substantial declines in the circulation of influenza and other respiratory diseases such as respiratory syncytial virus (RSV) raised questions about how to interpret existing and new surveillance metrics in the presence of COVID.^5,6^ Given these changes, along with changes in reporting practices during COVID-19 and increased public health concern about overall hospital capacity, the CDC switched the influenza forecasting target from percent ILI to weekly influenza hospital admissions beginning in 2022.^3,7^

These newly available data sources and COVID-19 driven changes in influenza transmission patterns, healthcare seeking behavior, and reporting requirements provide a unique opportunity to assess correlations across existing and new influenza surveillance data sources. Although comprehensive analysis of comparative COVID-19 signals has been undertaken,^8^ similar comparisons have not been conducted for influenza surveillance systems in the wake of COVID-19. These surveillance systems provide different measures of influenza activity, severity, and burden, and understanding how they correlate provides insight into the overall picture provided by influenza surveillance.

The state of California is a diverse and populous state with around 40 million residents. Influenza surveillance data sources for the state of California differ in their dates of availability, geographical and temporal resolution, and reporting frequency (Table 1).^9^ For influenza in the state of California, we: (1) explore how well different syndromic and sentinel surveillance data sources correlate with NHSN hospitalization admissions data; (2) explore which signals could serve as a potential leading input for forecasting NHSN hospital admissions; and (3) where possible with available data coverage and resolution, we assess spatiotemporal heterogeneity in these comparisons, especially differences across seasons and regions. We use NHSN influenza admissions as the reference metric because it is a target metric for influenza forecasting^10^ and because hospital capacity is of critical public health concern during winter respiratory virus seasons.

**Table 1.** Data sources included in the analysis with details about target, dates of availability, cadence, geographic coverage and resolution, reporting frequency and reporting lag.

| Target | Data source | Dates of availability and temporal resolution | Geographic coverage and resolution | Reporting frequency; lag |
| --- | --- | --- | --- | --- |
| **Percent positive influenza tests (%)** | Clinical laboratory | 2009 to present; weekly | 14 clinical labs during the analysis period | Weekly; 1-2 weeks reporting lag |
| **Number of influenza deaths** | Death certificates (text, coded, or other significant condition) | 2019 - /present; weekly | All of California; county-level | Weekly; 3-4 weeks reporting lag |
| **Number of positive influenza laboratory tests** | Electronic laboratory reporting (ELR) | From October 1, 2019; Mandated reporting of negatives began June 2023 with Title 17; daily | All of California, excluding LA for regional analysis; patient-level | Daily reporting frequency |
| **Influenza hospital admissions (non-laboratory confirmed)** | Department of Health Care Access and Information (HCAI) | 2000 - 2023 | All of California; patient-level | Daily resolution; 1-2 years lagged in reporting |
| **Percent ILI patients (%)** | Influenza like illness (ILI) | 2009 - present; Weekly | 48 counties | Weekly; 1-2 weeks lag |
| **Laboratory-confirmed influenza hospital admissions** | National healthcare safety network (NHSN) | February 2, 2022- April 30, 2024; Nov 1, 2024-present; daily prior to Nov. 2024; weekly thereafter | All of California; facility-level | Weekly; for 2023-2024 season, up to 10 days reporting lag |
| **Percent of emergency department visits that had a discharge diagnosis code of influenza (%)** | National Syndromic Surveillance Program (NSSP) | October 1, 2022 to present | State-level resolution; levels of enrollment and coverage have changed over time | Weekly |
| **Wastewater viral activity level (WVAL); influenza A subtype** | CDC wastewater (WW) surveillance | January 2022 to present; weekly | State level resolution only | Weekly |

## Methods

### Data sources

#### NHSN laboratory-confirmed hospital admissions

The Centers for Medicare and Medicaid Services (CMS) required all affiliated hospitals to report daily influenza information via the HHS Patient Impact and Hospital Capacity Data System beginning February 2, 2022.^7^ After the end of the public health emergency, NHSN entered a voluntary reporting period between April 30, 2024 and October 31, 2024.^11^ Mandated reporting of these data resumed November 1, 2024.^12^ Although this voluntary reporting period did not coincide with peak influenza activity, we estimated admissions under mandatory reporting by upscaling the data for this period based on the proportion of historical burden was contributed by those facilities that reported voluntarily.

#### Department of Health Care Access and Information (HCAI) hospital admissions

HCAI includes individual-level patient discharge data from across the state of California. Time series of influenza hospital admissions were compiled based on ICD-10 coding and date of admission. However, these data have a substantial availability delay of a year or more and are not available in real-time for ongoing surveillance.

#### National Syndromic Surveillance Program (NSSP) emergency department (ED) visits

Data from the NSSP was sourced from the CDC.^13^ This dataset became available for California beginning October 22, 2022, but notably, ED enrollment in California remained well below 50% during the analysis period.

#### Wastewater (WW) surveillance

State-level wastewater viral activity level (WVAL) data are sourced from the National Wastewater Surveillance System (NWSS).^14^ California data in NWSS included Influenza A wastewater concentrations measured by three programs, including the California Department of Public Health (CDPH) Drinking Water and Radiation Laboratory,^15^ the WastewaterSCAN program,^16,17^ and the NWSS commercial wastewater monitoring contract.^18^ All three programs used the same primers and probes,^19^ until November 25, 2024 when the NWSS commercial wastewater monitoring contract and WastewaterSCAN added an additional set of primers.^20^ The use of WVAL aims to harmonize differences in underlying wastewater sewershed systems and testing practices, but this metric is not currently calculated at a sub- state level, so regional WVAL data was not available for this analysis. Sewersheds covering approximately 60% of the California population are included in the WVAL metric. However, sewershed inclusion has varied substantially during the study period, resulting in inconsistent geographic and population coverage. Population-level sewershed coverage increased from about 10% at the beginning of 2022 to about 65% at the beginning of 2023. Since the beginning of 2023, the coverage has fluctuated between 55% and 65% of the California population. See Supplemental Notes for further discussion of potential limitations of the WW methodology.

#### Death certificate data

Weekly year-to-date dynamic death data are sourced from the CDPH Center for Health Statistics and Informatics. Influenza-coded deaths are any deaths with influenza listed in any immediate, contributing, or underlying cause of death field (whether specified as literal text or ICD-10 codes) on the death certificate. These influenza-coded deaths are not necessarily laboratory-confirmed and are an underestimate of all influenza-associated deaths.^21^

#### Clinical laboratory surveillance

Clinical sentinel laboratories report laboratory-confirmed influenza tests to CDPH on a weekly basis. These data have been used historically for monitoring the percentage test positivity.^9^ One geographic limitation for this data source is that typically only the location of the lab that submitted the specimen is known and not necessarily the county of residence of the patient.

#### Electronic laboratory reporting

Electronic laboratory reporting (ELR) data represent individual, patient level results statewide and were first available in October 2019 when positive influenza results first became reportable to public health.^22^ Required reporting of non-positive influenza results began in June 2023. Los Angeles results are excluded from region-specific analyses because these regional results are not yet fully integrated into reporting.

#### Influenza like illness (ILI)

ILI reporting comes from CDC sentinel medical providers throughout the state. Providers report the weekly number of outpatient visits attributable to ILI, which is defined as any patient presenting with a fever (> 100°F [37.8°C]) and cough or sore throat.^21,23^

### Analysis

We classified respiratory virus seasons following the standard definitions for Morbidity and Mortality Weekly Report (MMWR) influenza surveillance seasons, which extend from week 40 of a calendar year to week 39 of the following calendar year. Our analysis period began with the start date of mandatory reporting of laboratory-confirmed influenza hospitalizations on February 2, 2022 and continued through May 31, 2025. All data sources were aggregated to a standard MMWR week (i.e., Sunday through Saturday) for consistency.

Trends were assessed at a state level and across six public health regions. California public health regions were delineated during the COVID-19 pandemic through collaboration with CDPH and local health jurisdictions (Figure 1C). The six regions are groupings of city and county health jurisdictions designated as Rural North, Greater Sierra Sacramento, Bay Area, Central California, Southern California and Los Angeles.^24^ Notably, the Rural North is a less populous region that has less robust surveillance data.

**Figure 1.**
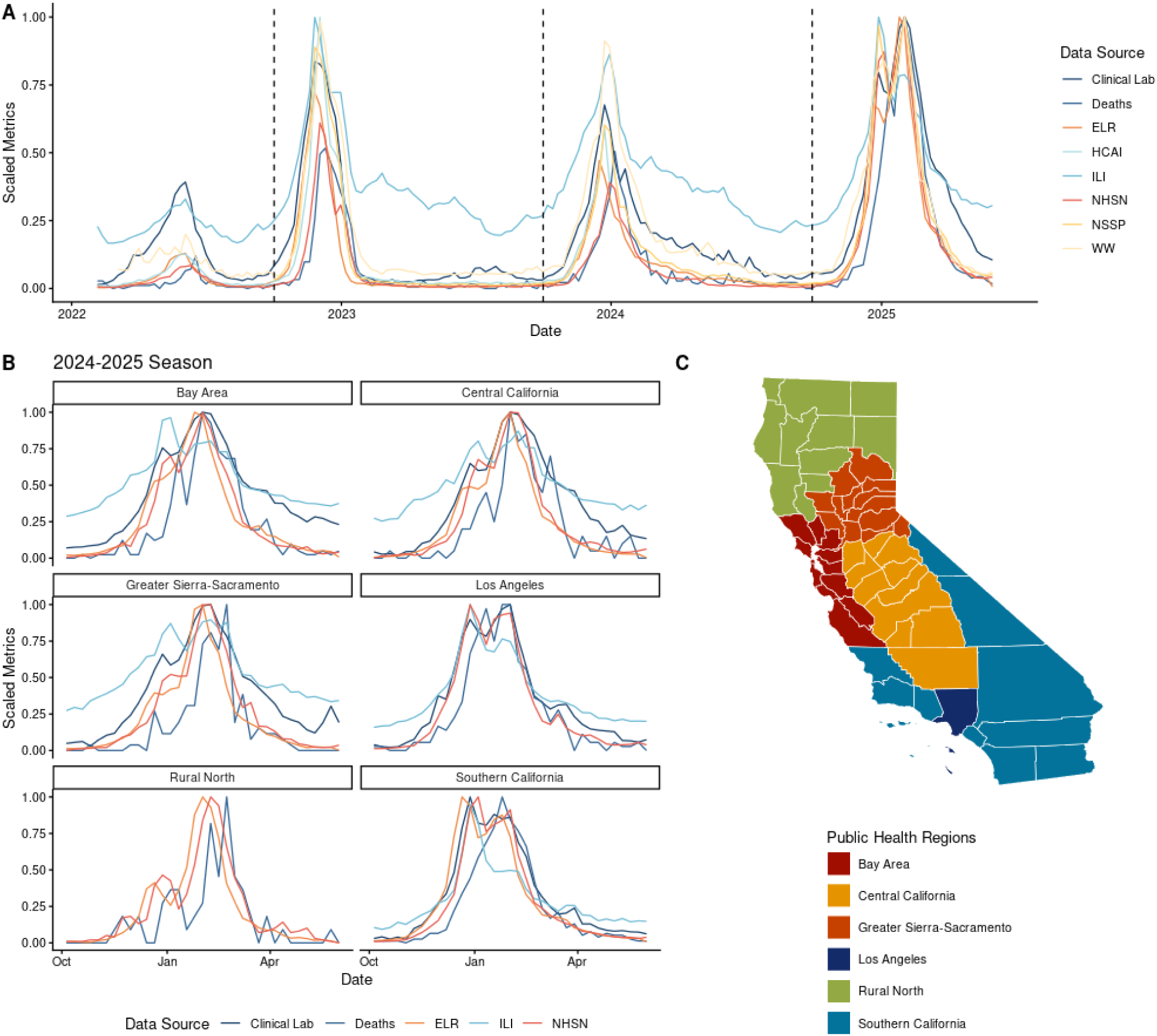
Scaled time course of various influenza surveillance data sources vs. NHSN laboratory-confirmed influenza admissions for (A) the state of California and (B) its public health regions. from February 2, 2022- May 1, 2025. (A) Time series of various influenza surveillance data sources vs. NHSN laboratory-confirmed influenza admissions for the state of California. Each signal is scaled to its maximum value within each respective region during the entire February 2, 2022- May 1, 2025 analysis period. Vertical dashed lines correspond to transitions between MMWR respiratory virus seasons. Note: Due to reporting delays, HCAI is not available after January 1, 2024, and so relative magnitude comparisons with respect to HCAI should be interpreted with caution. (B) Time series of various influenza surveillance data sources vs. NHSN laboratory-confirmed influenza admissions for the six public health regions of California for the most recent 2024-2025 MMWR season. (C) County level map of the public health regions within California. Acronyms: “Clinical Lab”= Clinical Laboratory; “Deaths”= death certificates; “ELR” = electronic laboratory reporting; “HCAI”= Department of Health Care Access and Information; “ILI”= influenza like illness; “NHSN”= National Healthcare Safety Network; “NSSP”= National Syndromic Surveillance Program; “WW”= Wastewater. Descriptions of each data source are available in Table 1.

To assess correlation with NHSN data, we scaled each data source in Table 1 to its maximum value during the entire February 2, 2022- May 31, 2025 period. We plotted each data source for visual inspection to compare across seasons and public health regions of California. We also generated scatterplots where the scaled data source was plotted against scaled NHSN admissions and compared to a 1:1 slope line. To quantify these correlations, we fit linear models of the scaled data sources as a predictor for scaled NHSN hospital admissions, fixing the y-intercept to zero.

As some data sources displayed dispersion and non-linearities, we also calculated Spearman’s correlation for each data source vs. NHSN hospital admissions for each available season and for the complete analysis period. We further calculated the cross correlation of each data source with respect to NHSN hospital admissions to explore which signals, if any, serve as a potential leading indicator for NHSN hospital admissions (i.e., exhibit trends that precede those same patterns in hospital admissions). Signals were compared at weekly lag intervals. Lastly, we conducted a principal component analysis (PCA) to understand how much variance across these different data sources could be explained by a single axis of dimensionality (i.e., exploring how much data sources overlap in the information they provide). Where possible with available data coverage and resolution, we assess spatiotemporal heterogeneity in these comparisons, especially differences by season and across regions of California.

All analyses were conducted in R version 4.2.3.^25^ Spearman’s correlation and cross correlation values were calculated using the “cor” and “ccf” functions respectively in the base stats package. PCA analysis was conducted using the “prcomp” function in the base stats package. Input values were scaled to unit variance prior to conducting the PCA analysis.

## Results

There was minimal influenza activity in the 2020-2021 and 2021-2022 flu seasons in California during the early COVID-19 pandemic. Influenza activity rebounded with a small, off-season peak of hospitalizations in summer 2022 and an earlier than expected start to the season in fall 2023; influenza activity resumed a more typical seasonal pattern in subsequent seasons (Figure 1A). However, different data sources portrayed the relative magnitudes and temporal progression of these three post-COVID seasons differently (Figure 1A), and there was also substantial regional variation (Figure 1B, Supplemental Figure 1). For example, while all data sources indicate a double peak for the 2024-2025 season, the relative magnitude and timing of those peaks differ by surveillance source (Figure 1A). Scaled ILI and wastewater signals peaked at higher levels statewide than NHSN admissions in 2022-2023 and 2023- 2024 seasons, and scaled ILI remained elevated across most regions during summer periods when other signals generally subsided (Figure 1, Supplementary Figure 1).

Linear regression results for scaled NHSN admissions vs. scaled alternative influenza data sources yielded slope estimates ranging from 0.4 (ILI) to > 0.9 (deaths and ELR) (Supplementary Table 1 & Supplementary Figure 2A). There was also variation in these slope estimates across regions (Supplementary Figure 2B).

### Spearman’s correlation

In general, all influenza surveillance data sources in California had high Spearman’s correlation with NHSN laboratory-confirmed influenza hospitalizations (ρ ≥ 0.80) (Figure 2, Table 2). These relationships generally strengthened through time from the 2021-2022 season to the most recent 2024-2025 season (Supplementary Figure 3). Deaths, ILI, clinical lab, and WW showed a notable improvement in correlation strength between the 2021-2022 and the most recent 2024-2025 seasons. ELR, HCAI, and NSSP data had the strongest Spearman’s correlations for the entire analysis period (ρ ≥ 0.90), while deaths and ILI had lower correlation strengths (ρ = 0.84 and 0.80 respectively) (Figure 2, Table 2).

**Figure 2.**
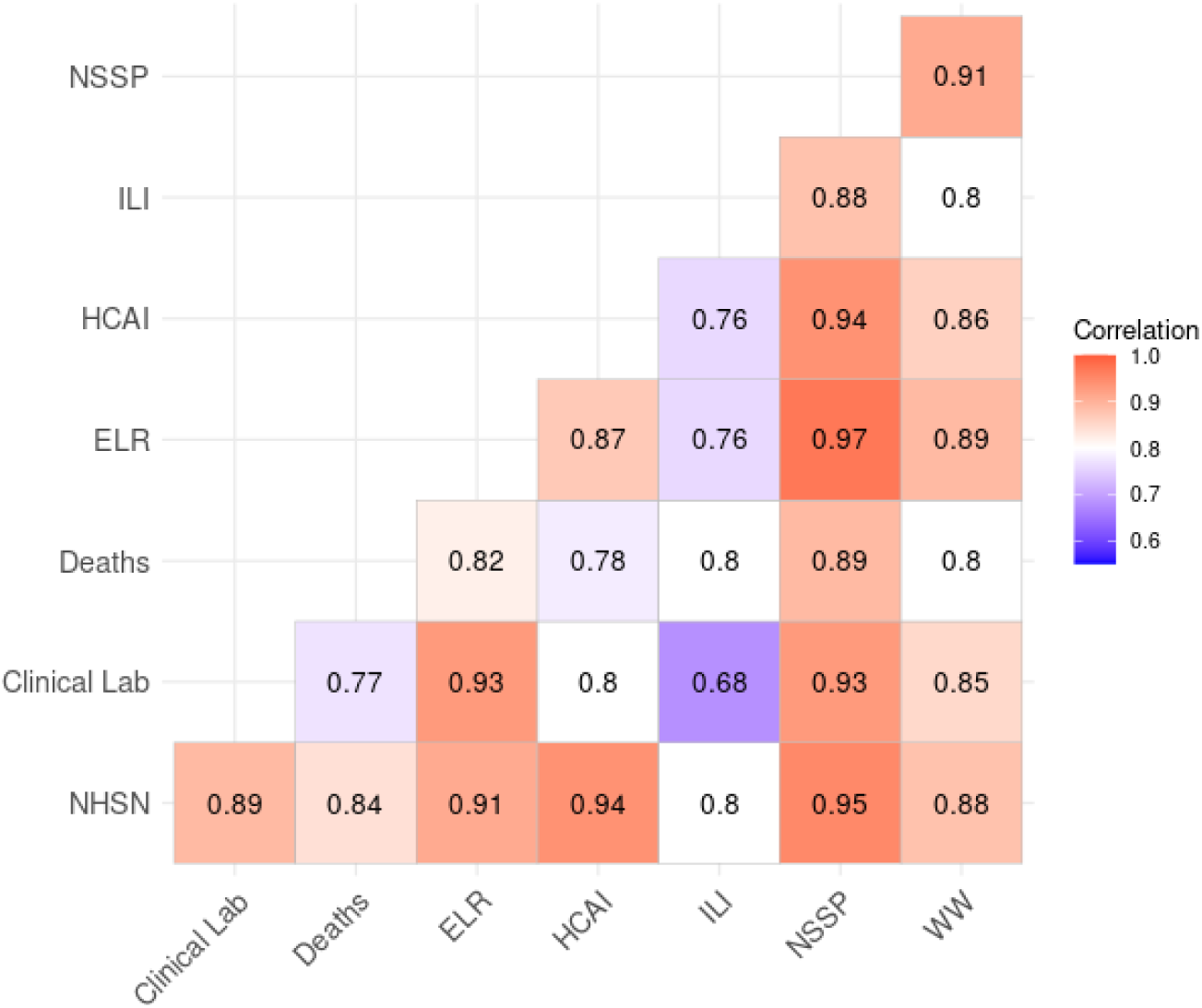
Correlation heat map of influenza data sources for all seasons from 2022-2025. Values in heat map correspond to Spearman’s correlation values. Values closer to one indicate stronger correlation. Acronyms: “Clinical Lab”= Clinical Laboratory; “Deaths”= death certificates; “ELR” = electronic laboratory reporting; “HCAI”= Department of Health Care Access and Information; “ILI”= influenza like illness; “NHSN”= National Healthcare Safety Network; “NSSP”= National Syndromic Surveillance Program; “WW”= Wastewater. Descriptions of each data source are available in Table 1.

**Table 2.**
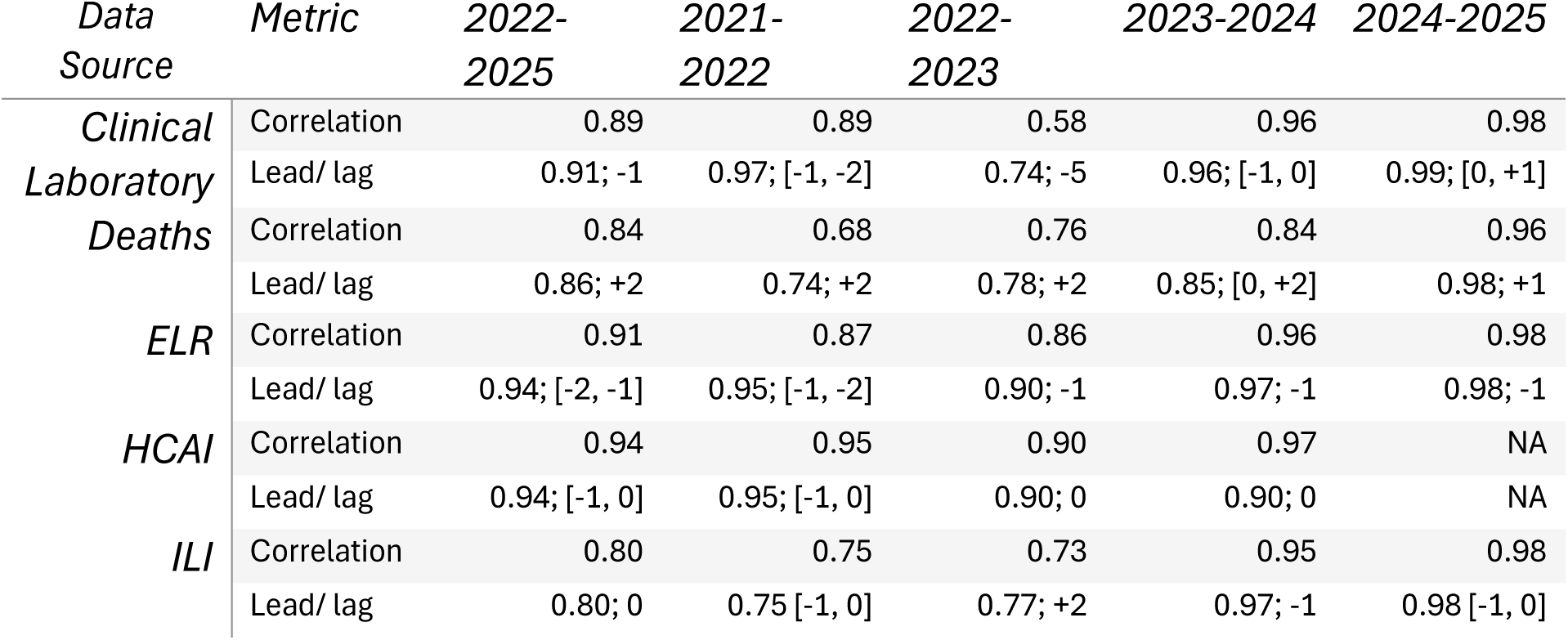
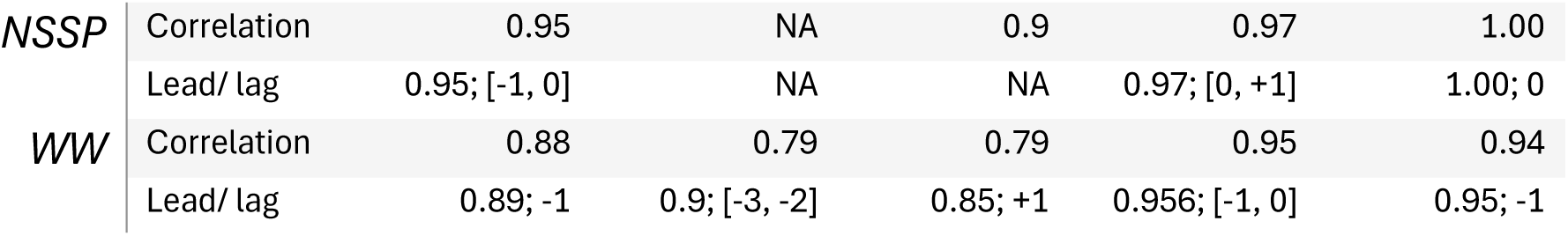
Summary of Spearman’s correlation and cross correlation results for different influenza surveillance data sources relative to NHSN lab-confirmed hospital admissions across individual seasons and the entire analysis period. Due to reporting delays, HCAI is not available after January 1, 2024, and so the 2023-2024 season reflects correlation for only the available data prior to the start of the new year. Acronyms: “Deaths”= death certificates; “ELR” = electronic laboratory reporting; “HCAI”=Department of Health Care Access and Information; “ILI”= influenza like illness; “NHSN”= National Healthcare Safety Network; “NSSP”= National Syndromic Surveillance Program; “WW”= Wastewater. Descriptions of each data source are available in Table 1.

For the entire analysis period, there was also variation in correlation strength between different signals across public health regions (Supplementary Figure 4). Deaths also had lower correlation values compared to all other surveillance signals in the Rural North (ρ ≤ 0.58). Of available signals at the regional scale, HCAI (ρ = [0.82, 0.92]) and ELR (ρ = [0.83, 0.87]) data generally had the highest Spearman’s correlation values with NHSN data (Supplementary Figure 4).

### Cross correlation

ELR was the only data source with the highest correlation values at one to two weeks ahead of NHSN admissions across all four seasons statewide—with Spearman’s ρ ≥ 0.90 across all four seasons for optimal lags (Figure 3). Clinical laboratory data consistently led NHSN time series (negative lag values) across seasons and regions, except for the 2024-2025 season. Apart from deaths which consistently lagged NHSN hospital admissions by one to two weeks, most data signals neither consistently led nor lagged NHSN hospital admissions across all four seasons and regions (Figure 3, Supplementary Figure 5). Even though wastewater and ILI had comparatively lower Spearman’s correlation values relative to some other data sources for the entire analysis period, they still potentially served as leading signals in the 2021-2022, 2023-2024, and 2024-2025 seasons.

**Figure 3.**
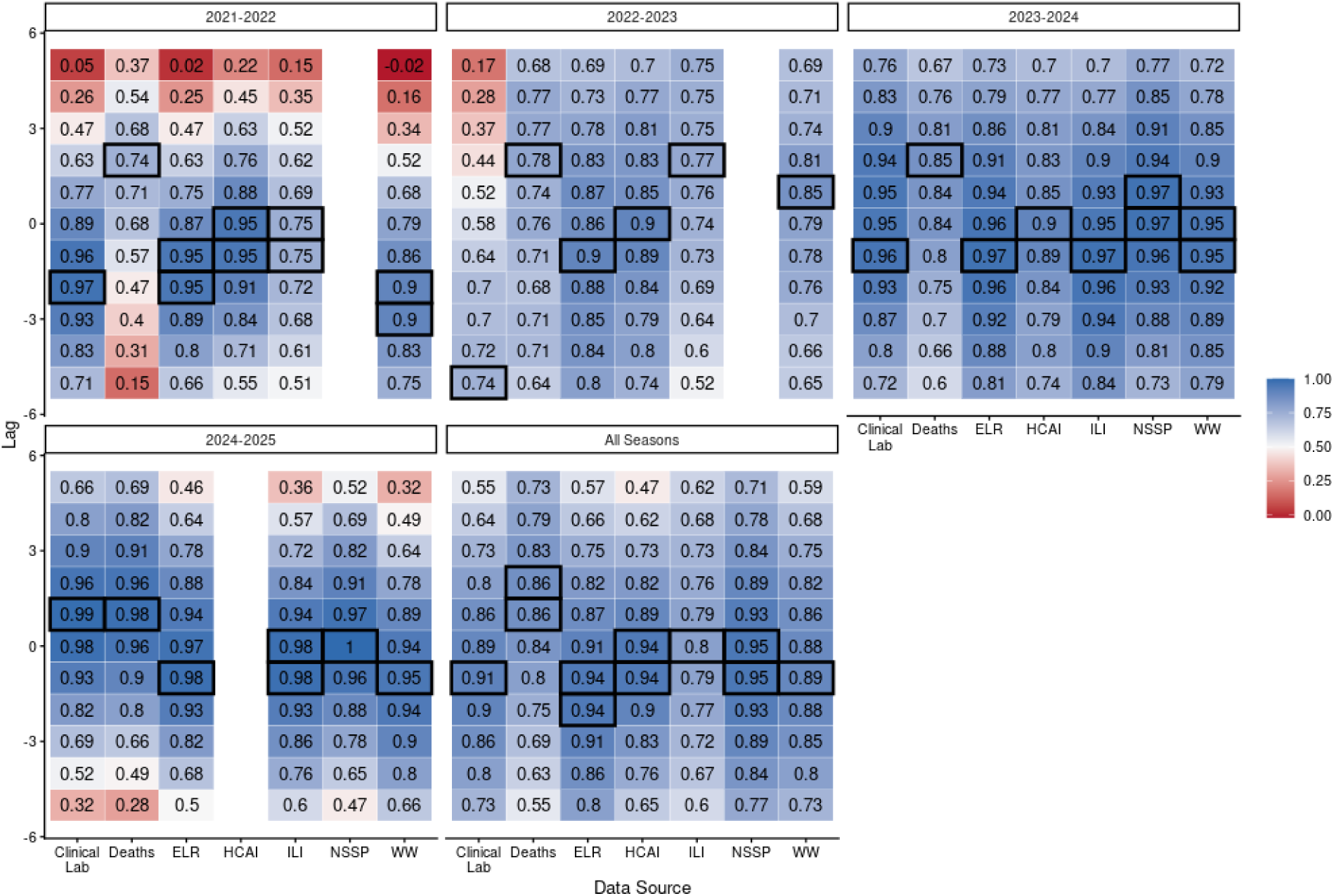
Heat map showing cross correlation values between various influenza data signals and NHSN laboratory-confirmed influenza admissions across different seasons for the state of California. Lag units on the y-axis correspond to weeks. Correlation values in the figure correspond to Spearman’s correlation values. Strongly correlated negative lag values indicate a signal that potentially leads NHSN admissions, while strongly correlated positive lag values indicate a signal that potentially lags NHSN admission. Signals were compared at weekly lag intervals. The peak correlation value (rounded to two decimal places) is highlighted for each signal with a black outline box. Acronyms: “Clinical Lab”= Clinical Laboratory; “Deaths”= death certificates; “ELR” = electronic laboratory reporting; “HCAI”= Department of Health Care Access and Information; “ILI”= influenza like illness; “NHSN”= National Healthcare Safety Network; “NSSP”= National Syndromic Surveillance Program; “WW”= Wastewater. Descriptions of each data source are available in Table 1.

### Principal Component Analysis (PCA)

PCA analysis suggests that 93% of the variation in data signals can be explained by a single principal component. For the second component, which explains only 4% of the variation, deaths and ILI were the most diverse in their correlation strengths (Figure 4, Supplementary Table 2). All variables had a similar correlation strength with the first principal component ranging from 0.36 to 0.39 (Supplementary Table 2). The second principal component exhibited more variation with correlation strengths ranging from - 0.62 for ILI to 0.63 for deaths (Supplementary Table 2).

**Figure 4.**
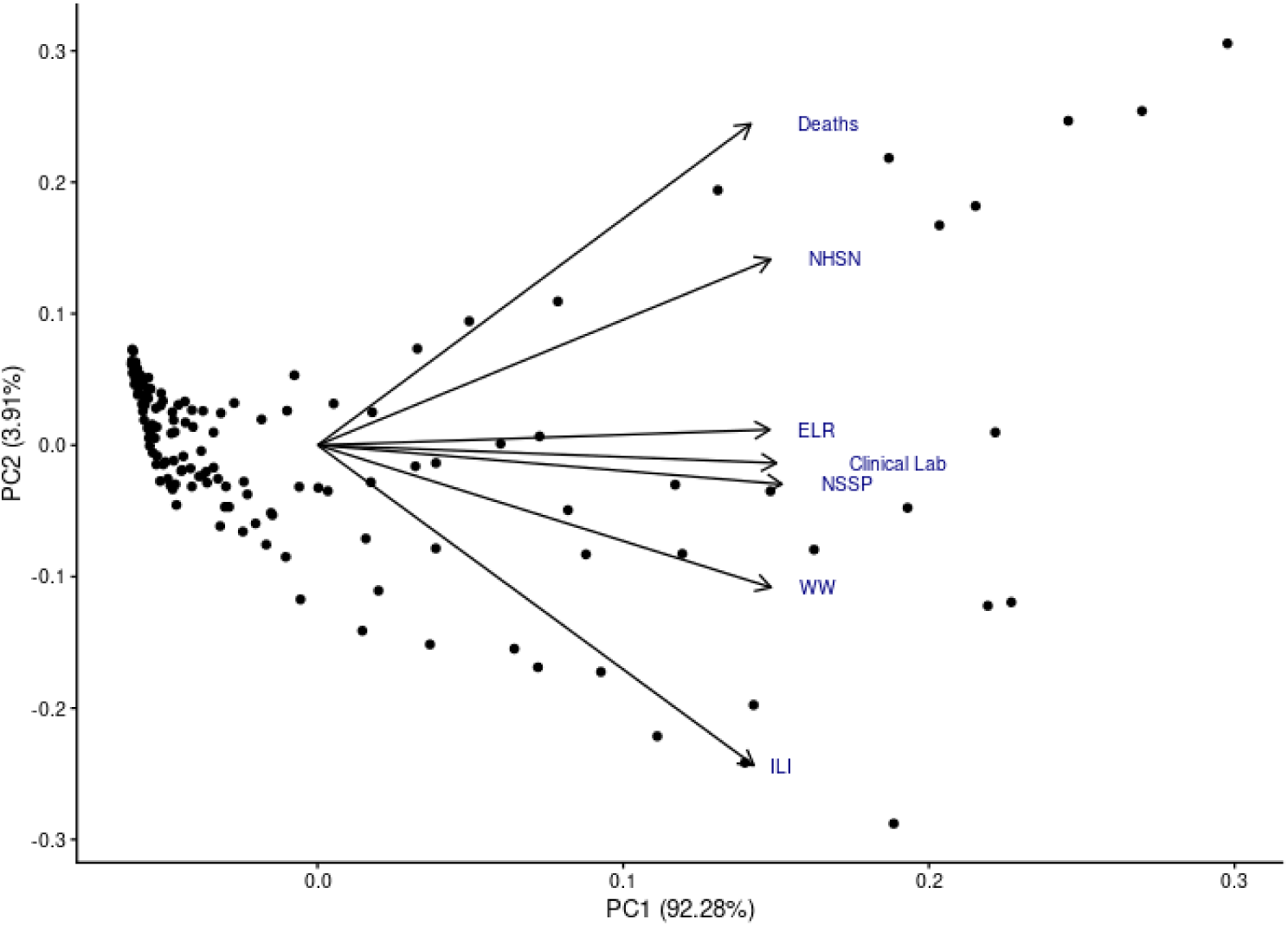
Principal component analysis results. Each point corresponds to a single MMWR reporting week. Acronyms: “Clinical Lab”= Clinical Laboratory; “Deaths”= death certificates; “ELR” = electronic laboratory reporting; “HCAI”=Department of Health Care Access and Information; “ILI”= influenza like illness; “NHSN”= National Healthcare Safety Network; “NSSP”= National Syndromic Surveillance Program; “WW”= Wastewater. Descriptions of each data source are available in Table 1.

## Discussion

Newly mandated reporting of laboratory-confirmed hospital influenza admissions provided a new, population-based measure of burden. In this analysis, we compared historical data from syndromic and sentinel influenza surveillance systems to laboratory-confirmed NHSN influenza admissions across seasons and regions, with the goal of identifying strengths and weaknesses for potential analytical applications including surveillance, forecasting, and scenario modeling. COVID-19 starkly altered influenza transmission during the first few years of the pandemic; changes in testing patterns and potential conflation with COVID-19 further complicated the interpretation of traditional influenza surveillance data.^5^

The diverse data sources in this analysis (Table 1) all have potential tradeoffs in reporting delays (e.g., HCAI [≥1 year], deaths [3-4 weeks]), geographic coverage, resolution, and biases that reflect the diverse goals of influenza surveillance. For example, laboratory data (e.g., ELR and clinical laboratory) and outpatient visits (e.g., NSSP, ILI), are more focused on identifying the onset of the season and season intensity, while others like hospitalizations and deaths also serve to measure season severity.

Nonetheless, although patterns for individual signals varied across seasons and regions (Figures 1, Supplementary Figure 1), there was a high Spearman’s correlation (ρ ≥ 0.8) between all data sources and NHSN data (Figure 2).

Indeed, the most aligned datasets (NSSP, clinical laboratory, HCAI, and ELR data) all exceeded Spearman’s correlation ρ ≥ 0.90 for the complete analysis period. In addition, the large amount of variance explained by a single PCA axis (Figure 4), supports the idea that these data sources are generally quite congruent. However, many of the signals in this analysis started with lower correlation values in the 2021-2022 season and continued to improve through the most recent 2024-2025 season. This was especially notable for ILI, deaths, ELR, clinical lab, and WW datasets (Supplementary Figure 3). This pattern may reflect a “there and back again” story with the return of more typical influenza seasonal patterns, clinician awareness and diagnostic capacity to differentiate between COVID-19 and influenza, and improvements in coverage or reporting completeness in comparatively newer surveillance sources like ELR and WW.

From an early warning or leading indicator perspective, there was variability in whether signals led or lagged NHSN admissions when comparing across seasons and regions of California (Figure 3, Supplementary Figure 5, Table 2). While NSSP had highest overall Spearman’s correlation of all data sources with NHSN data, it was not a leading indicator as measured with a weekly time step. ELR and clinical laboratory data provided the most consistent leading signal, which could be because these are the only surveillance data sources that also measure laboratory-confirmed influenza. However, these two signals currently lack geographic coverage for the state in Los Angeles and Rural North regions, respectively. In the context of implementing forecasting or predictive analytics, the one-to-two-week delay in clinical laboratory data reporting also potentially limits its utility for short-term forecasting.

Other signals like ILI and WW provided an early warning signal relative to NHSN (sometimes up to three weeks an advance), but since this was not consistent across seasons, this makes confidence in its application for predictive analytics more challenging. Since we examined cross correlations with time steps of weeks, it may be possible that other syndromic signals like NSSP ED visits could still provide a few days leading signal, but we lacked the temporal resolution to assess this adequately.

These analyses were designed to lend insight into the relationships between different influenza surveillance data sources and lab-confirmed hospitalizations in California, including by region and across seasons. While many of these data sources are available in other states, our results are not necessarily generalizable to other locations given the potential differences in population-level coverage or implementation. Several data sources (i.e., WW and NSSP) had changing patterns of coverage or had limitations in regional availability during the analysis period, limiting full exploration of regional differences. We have also treated NHSN as the primary comparator and used it without modification except for imputation during the voluntary reporting period. Nevertheless, the number of facilities reporting to NHSN week-by-week fluctuates over time. It is also important to note that patients captured by laboratory-confirmed data may have different demographics than those represented in syndromic surveillance. Lastly, from a predictive analytics perspective, this analysis does not account for any backfilling that likely occurs in real-time reporting; those potential differences may be important to consider for forecasting applications.

Since the beginning of the COVID-19 pandemic, the HHS Protect/NHSN data grew in importance for public health by providing real-time information about hospital admissions and census at a time when hospital capacity was a critical indicator. This analysis demonstrates that pre-existing influenza surveillance and sentinel data sources correspond well with this new signal, and that this agreement has generally improved over time. In the meantime, NHSN continues to offer real-time reporting at the state and facility levels that will remain critical in surveillance of future respiratory virus seasons across the United States.

## Acknowledgements

The authors would like to thank Gail Sondermeyer-Cooksey for assistance with extracting HCAI data and Alexander Yu for constructive comments on the manuscript.

## Data availability

Code and scaled data to reproduce the analyses are available at: https://github.com/cdphmodeling/flu_correlations.

NSSP data at the state-level are publicly available at: https://data.cdc.gov/Public-Health-Surveillance/NSSP-Emergency-Department-Visit-Trajectories-by-St/rdmq-nq56/about_data

NHSN data are publicly available at: https://healthdata.gov/dataset/Weekly-United-States-Hospitalization-Metrics-by-Ju/kruc-9unj/about_data and https://data.cdc.gov/Public-Health-Surveillance/Weekly-Hospital-Respiratory-Data-HRD-Metrics-by-Ju/ua7e-t2fy/about_data

State-level wastewater data are available from the CDC at: https://www.cdc.gov/nwss/rv/InfluenzaA-statetrend.html

Syndromic influenza hospitalization data derived from the California Department of Healthcare Access and Information (HCAI) are available upon a data request: https://hcai.ca.gov/data/request-data/

## Ethics

The California Health and Human Services Agency Committee for the Protection of Human Subjects (CPHS) has determined that this research (project number 2024-210) is classified as exempt under the federal Common Rule. This decision is issued under the California Health and Human Services Agency’s Federalwide Assurance #00000681 with the Office of Human Research Protections (OHRP).

## Authors contributions

LW: conceptualization, formal analysis, methodology, validation, visualization, writing—original draft, writing—review and editing; MS, EB, CH, CP, NL, FC, CS: data curation, writing—review and editing; EM: conceptualization, data curation, writing—review and editing; TL: conceptualization, methodology, supervision, writing—original draft, writing—review and editing.

## Conflict of interest declaration

The authors declare no competing interests.

## Funding

This work was supported by the California Department of Public Health. The findings and conclusions in this article are those of the author(s) and do not necessarily represent the views or opinions of the California Department of Public Health or the California Health and Human Services Agency.

This study used the California Patient Discharge Dataset. The interpretation and reporting of these data are the sole responsibility of the authors. The authors acknowledge the California Department of Healthcare Access and Information for compilation of these data.

This work was funded by Centers for Disease Control and Prevention, Epidemiology and Laboratory Capacity for Infectious Diseases, Cooperative Agreement Number 6 NU50CK000539.

## Notes

### Competing Interest Statement

The authors have declared no competing interest.

