## Supplementary for "A new surveillance landscape for seasonal influenza?: Comparing lab-confirmed influenza hospitalizations with other syndromic and surveillance data sources for the state of California"

**Supplementary Materials**

**Supplemental Notes on Wastewater Methods Limitations:** The wastewater WVAL utilized in this manuscript is a publicly available tool, published by the CDC NWSS team and available for download. Because the WVAL is continuously updated on the CDC's webpage, the exact version of the WVAL used in this study is available in the code repository associated with this paper. Multiple wastewater testing programs have contributed wastewater influenza A measurements to the CDC NWSS program's dataset, which constitutes the aggregated WVAL. The laboratory methods associated with these testing programs have been referenced in the text wherever possible. However, some contributing programs – including the CDPH State DWRL and the CDC NWSS wastewater testing contract – do not currently have published laboratory methods available for reference. Wastewater viral concentrations are microbiological measurements of environmental samples, where final measured concentrations arise from a series of steps, including sample collection, pre-processing, concentration, extraction, and quantitative measurement. There are multiple sources of biases and variation that may arise at every processing step, both from methodological choices and natural environmental variability. Given this high degree of potential bias and variability, it is important that detailed laboratory methods, quality control criteria, and quality control measurements be made available for published environmental microbiological data (https://doi.org/10.1021/acs.est.1c01767). This is especially true when data is intended to be used for public health decision-making and policy. Wastewater data as a public health tool for respiratory virus surveillance is still relatively new. Publicizing the relevant methods for published wastewater data will help build the usability and the credibility of this tool. However, although all associated methods are not readily available at the time of this publication, we have chosen to evaluate the WVAL as a flu surveillance tool because it is already published on a national dashboard and is presumably being used by both the public and public health stakeholders for situational awareness and decision-making.

**Supplementary Table 1. Linear model fit results for scaled NHSN admissions vs. scaled alternative influenza data sources fit to the formula: scaled NHSN = m*(scaled data source).** Acronyms: "Deaths"= death certificates; "ELR" = electronic laboratory reporting; "HCAI"= Department of Health Care Access and Information; "ILI"= influenza like illness; "NHSN"= National Healthcare Safety Network; "NSSP"= National Syndromic Surveillance Program; "WW"= Wastewater. Descriptions of each data source are available in Table 1.

| *Data source* | *Estimate* | *Standard error* | *Statistic* | *p-value* |
|---|---|---|---|---|
| *Clinical Laboratory* | 0.64 | 0.02 | 30.03 | < 0.001 |
| *Deaths* | 0.99 | 0.02 | 42.84 | < 0.001 |
| *ELR* | 0.91 | 0.02 | 41.07 | < 0.001 |
| *HCAI* | 0.61 | 0.01 | 44.52 | < 0.001 |
| *ILI* | 0.39 | 0.03 | 14.94 | < 0.001 |
| *NSSP* | 0.79 | 0.02 | 42.87 | < 0.001 |
| *WW* | 0.65 | 0.02 | 29.71 | < 0.001 |

**Supplementary Table 2. Coefficients of the PCA.** Component weights or loading for each variable in the PCA correspond to correlation coefficients with each respective principal component. Acronyms: "Deaths"= death certificates; "ELR" = electronic laboratory reporting; "HCAI"= Department of Health Care Access and Information; "ILI"= influenza like illness; "NHSN"= National Healthcare Safety Network; "NSSP"= National Syndromic Surveillance Program; "WW"= Wastewater. Descriptions of each data source are available in Table 1.

| | PC1 | PC2 | PC3 | PC4 | PC5 | PC6 | PC7 |
|---|---|---|---|---|---|---|---|
| **CLINICAL LABORATORY** | 0.39 | -0.03 | -0.21 | -0.71 | -0.11 | 0.33 | 0.41 |
| **DEATHS** | 0.36 | 0.63 | 0.48 | -0.24 | -0.15 | -0.34 | -0.22 |
| **ELR** | 0.38 | 0.03 | -0.68 | 0.09 | 0.12 | -0.61 | -0.01 |
| **ILI** | 0.37 | -0.62 | 0.31 | 0.15 | -0.55 | -0.23 | 0.06 |
| **NHSN** | 0.38 | 0.36 | -0.03 | 0.63 | -0.10 | 0.36 | 0.43 |
| **NSSP** | 0.39 | -0.08 | -0.19 | 0.05 | -0.03 | 0.47 | -0.76 |
| **WW** | 0.38 | -0.28 | 0.37 | 0.03 | 0.80 | -0.02 | 0.09 |

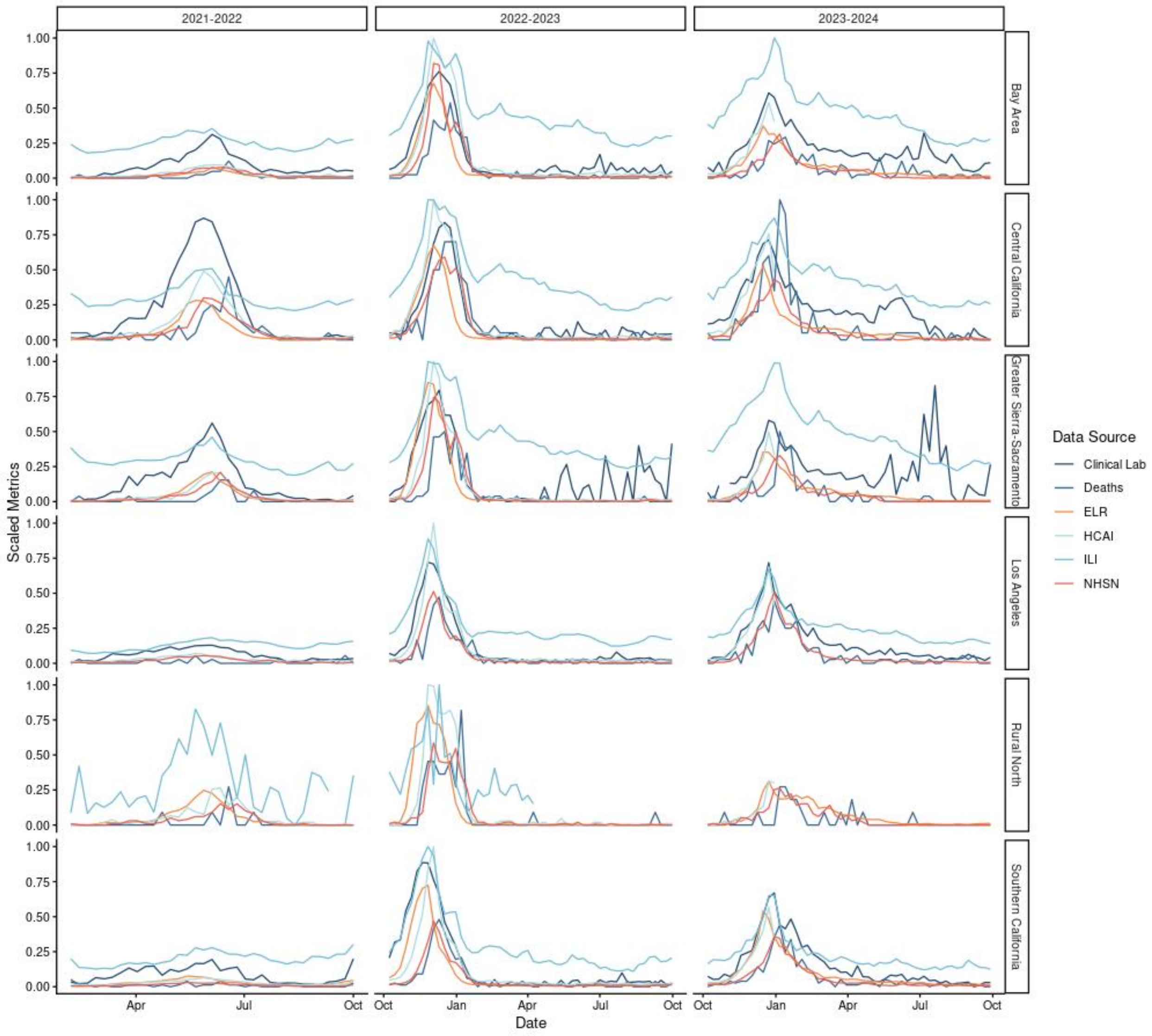


**Supplementary Figure 1. Time series of various influenza surveillance data sources vs. NHSN lab-confirmed influenza admissions for the six public health regions of California for the 2021-2022, 2022-2023, and 2023-2024 seasons.** Each signal is scaled to its maximum value within each respective region during the entire February 2, 2022- May 31, 2025 period. The Rural North region does not have clinical lab reporting, nor does it have ILI data available after April 8, 2023. Regional views for the 2024-2025 season are available in Figure 1B. Acronyms: “Clinical Lab”= Clinical Laboratory; “Deaths”= death certificates; “ELR” = electronic laboratory reporting; “HCAI”= Department of Health Care Access and Information; “ILI”= influenza like illness; “NHSN”= National Healthcare Safety Network. Descriptions of each data source are available in Table 1.

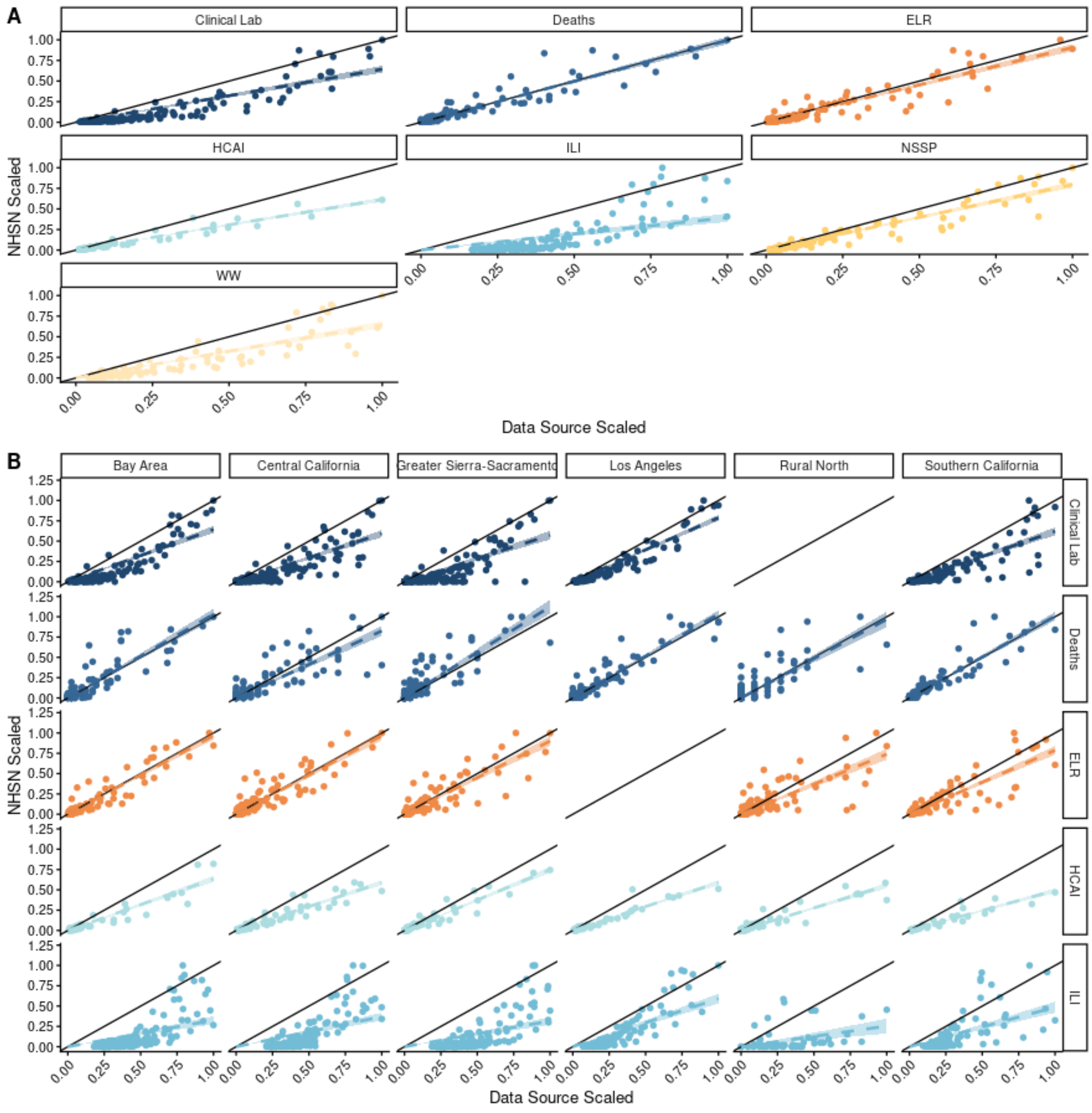


**Supplementary Figure 2. Scatter plots of various influenza data sources vs. NHSN lab-confirmed influenza admissions from February 2, 2022- May 1, 2025 for (A) the state of California and (B) six public health regions within the state of California.** Each plotted point represents the relationship between two data sources for a single epi week during the analysis period. The solid diagonal line represents a 1:1 relationship between NHSN admissions and the respective data sources. Acronyms: "Clinical Lab"= Clinical Laboratory; "Deaths"= death certificates; "ELR" = electronic laboratory reporting; "HCAI"= Department of Health Care Access and Information; "ILI"= influenza like illness; "NHSN"= National Healthcare Safety Network; "NSSP"= National Syndromic Surveillance Program; "WW"= Wastewater. Descriptions of each data source are available in Table 1.

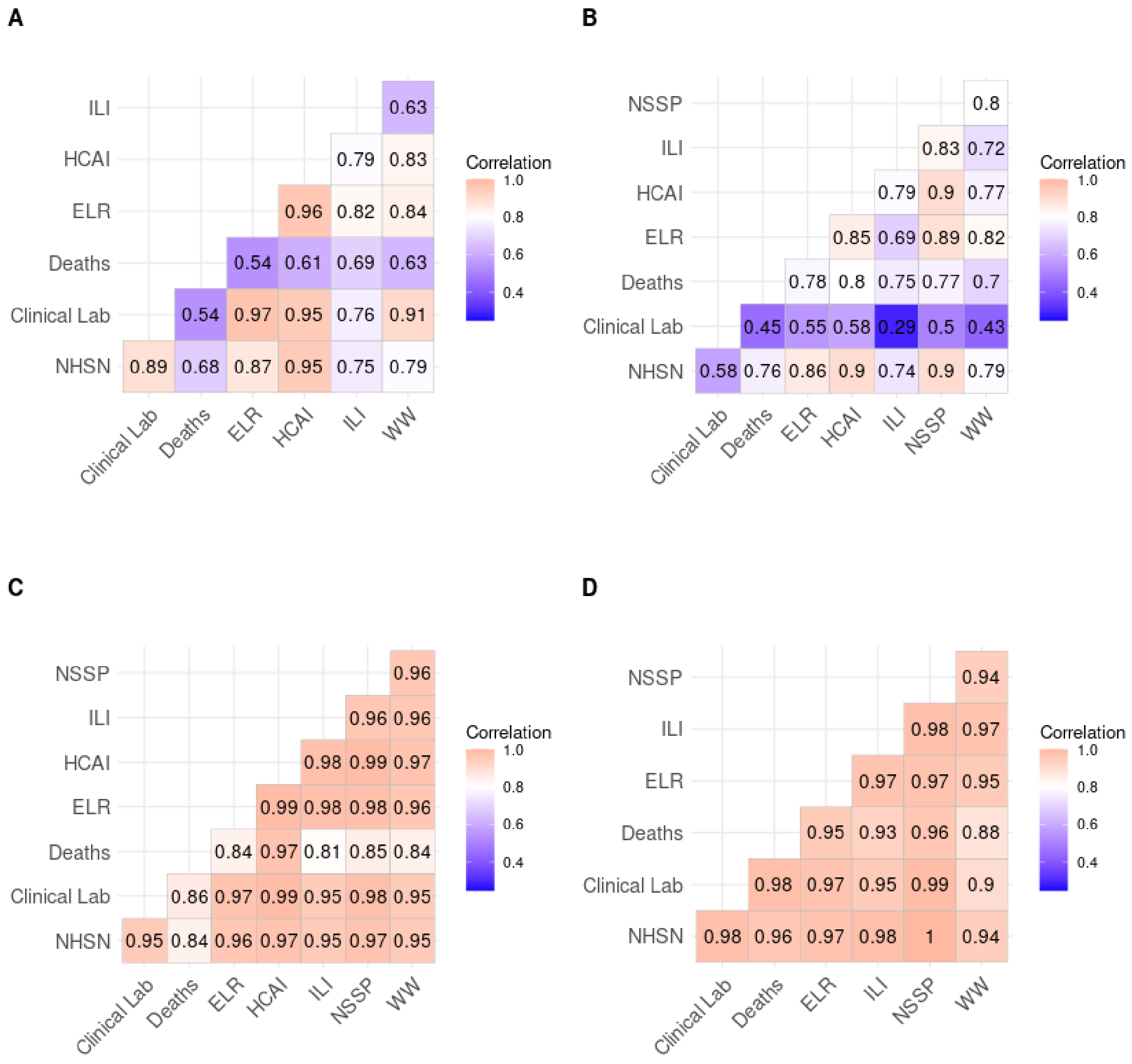


**Supplementary Figure 3. Correlation heat maps of influenza data sources for the state of California for individual respiratory virus seasons during the analysis period: (A) 2021-2022; (B) 2022-2023; (C) 2023-2024; (D) 2024-2025.** Values in heat map correspond to Spearman's correlation values. Values closer to one indicate stronger correlation. Acronyms: "Clinical Lab"=

Clinical Laboratory; “Deaths”= death certificates; “ELR” = electronic laboratory reporting; “HCAI”= Department of Health Care Access and Information; “ILI”= influenza like illness; “NHSN”= National Healthcare Safety Network; “WW”= Wastewater. Descriptions of each data source are available in Table 1.

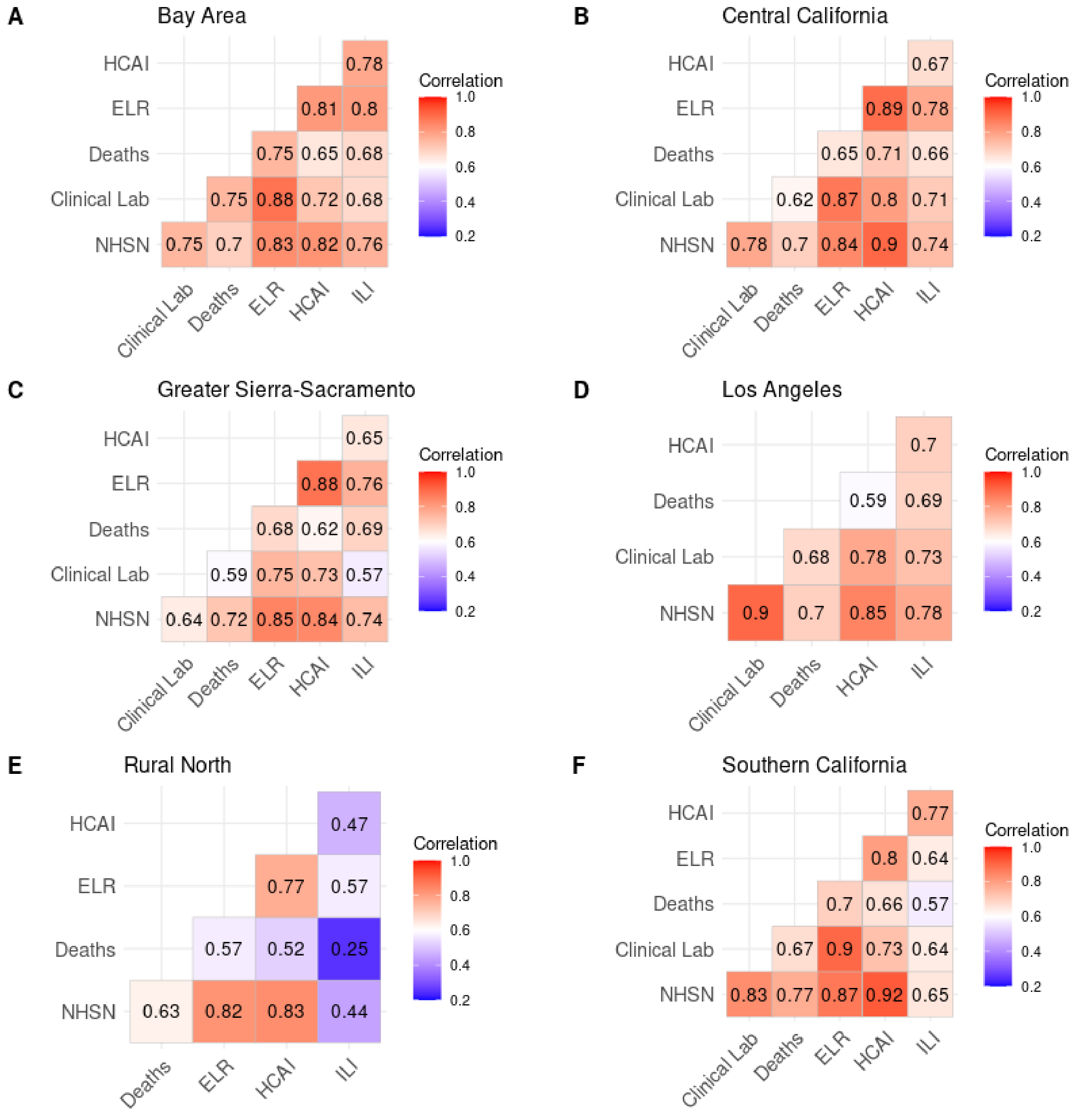

**Supplementary Figure 4. Correlation heat maps of influenza data sources for the six different public health regions of California from February 2, 2022- May 1, 2025.** Numerical values in heat map correspond to Spearman's correlation values. Values closer to one indicate stronger correlation. Acronyms: "Clinical Lab"= Clinical Laboratory; "Deaths"= death certificates; "ELR" = electronic laboratory reporting; "HCAI"= Department of Health Care Access and Information; "ILI"= influenza like illness; "NHSN"= National Healthcare Safety Network. Descriptions of each data source are available in Table 1.

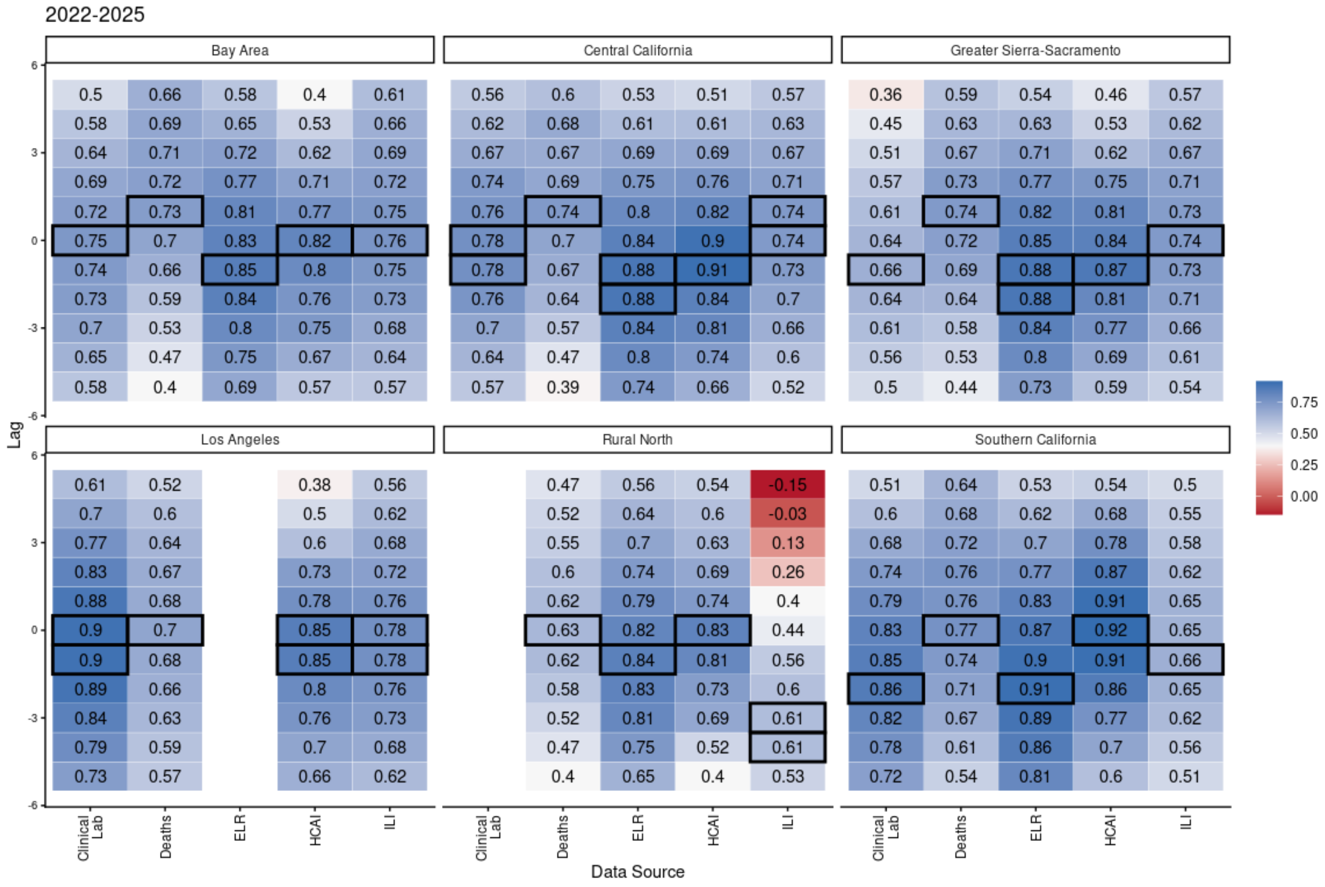


**Supplementary Figure 5. Heat map showing cross correlation between various influenza data signals and NHSN lab-confirmed influenza admissions across regions.** Strongly correlated negative lag values indicate a signal that potentially leads NHSN admissions, while positive lag signals indicate that the respective signal has a stronger correlation when lagging NHSN admissions. Acronyms: "Clinical Lab"= Clinical Laboratory; "Deaths"= death certificates; "ELR" = electronic laboratory reporting; "HCAI"= Department of Health Care Access and Information; "ILI"= influenza like illness; "NSSP"= National Syndromic Surveillance Program; "WW"= Wastewater. Descriptions of each data source are available in Table 1.
